# Calibrating CONSORT-AI with FAIR Principles to enhance reproducibility in AI-driven clinical trials

**DOI:** 10.1101/2025.07.07.25330987

**Authors:** Kirubel Biruk Shiferaw, Irina Balaur, Gary Collins, Curtis Sharma, Leyla Jael Castro, Fotis Psomopoulos, Daniel Garijo, Ron Henkel, Dagmar Waltemath, Atinkut Alamirrew Zeleke

## Abstract

Artificial intelligence (AI) is increasingly embedded in clinical trials, yet poor reproducibility remains a critical barrier to trustworthy and transparent research. In this study, we propose a structured calibration of the CONSORT-AI reporting guideline using the FAIR (Findable, Accessible, Interoperable, Reusable) principles. We introduce the application of CALIFRAME, a framework designed to evaluate and align existing medical AI reporting standards with FAIR-compliant practices. Applying CALIFRAME to the CONSORT-AI checklist reveals specific gaps in data and code sharing, metadata use, and accessibility practices in current AI-driven clinical trials. Our results underscore the need for standardized metadata, clear licensing, and stakeholder-inclusive design in medical AI reporting. We demonstrate that FAIR-oriented calibration of reporting guidelines can bridge the reproducibility gap and support more transparent, efficient, and reusable AI interventions in healthcare. This work advocates for a shift toward reproducibility as a foundation for trustworthy AI in clinical research.

## Introduction

The integration of Artificial Intelligence (AI) has significantly enhanced the potential of medical research, driving cutting-edge advancements in diagnostics, prognostics, and therapeutic interventions [1]. Recent studies indicated a growing number of Randomized Clinical Trials (RCTs) investigating the impact of AI driven interventions [2]. As the gold standard for efficacy and safety of medical interventions, RCTs play a crucial role in assessing the clinical utility of AI applications in healthcare [3].

In response to the increased number of clinical trials involving AI interventions, existing reporting guidelines for clinical trials have been updated to include essential reporting items specific to AI driven interventions [4]. Reporting guidelines are essential to ensure the transparency, reliability and reproducibility of research outcomes by providing a structured and comprehensive framework [5]. They facilitate clear communication among academic editors, reviewers, researchers and other stakeholders enabling a more rigorous appraisal of research findings. However, the integration of AI algorithms into the clinical domain requires not only the refinement of existing reporting guidelines but also the potential development of new frameworks that adequately address the unique challenges of AI based interventions.

The Consolidated Standards of Reporting Trials for Artificial Intelligence (CONSORT-AI) were introduced to enhance transparency and comprehensiveness in reporting clinical trials involving AI interventions [6]. However, a recent review of 65 RCTs conducted since 2020 found that only 10 adhered to CONSORT-AI [7]. Several key metadata items such as model or code accessibility, interoperability, licensing, algorithmic metadata (including version and dependencies), trial protocol, and reuse disclaimers were consistently under-reported.

Furthermore, only 3 out of the 52 reviewed journals explicitly endorsed CONSORT-AI as a mandatory reporting guideline for RCTs involving AI interventions [7]. While many journals may instead direct authors to the Enhancing the *QUAlity and Transparency Of health Research* (EQUATOR) Network for general guidance, the growing number of reporting guidelines presents a practical challenge making it difficult for journals to explicitly mandate or prioritize individual reporting guidelines. A related review focusing on RCTs with machine learning (ML) interventions revealed that, none of the 41 RCTs fully adhered to all the AI specific 14 items of CONSORT-AI checklist, and a considerable proportion 90% (37 studies) failed to disclose code availability [8]. Common reasons for nonadherence included a lack of performance error analysis and failure to assess the impact of unavailable/poor quality input data [8]. Despite the introduction of CONSORT-AI, key aspects critical to AI-driven research such as data and code accessibility remain inconsistently reported [8]. While CONSORT-AI importantly recommends that authors “state whether and how the AI intervention and/or its code can be accessed, including any restrictions to access or re-use”, it does not yet offer detailed guidance on how to report technical metadata such as model architecture, training procedures, software dependencies, or licensing. This lack of structured detail may limit reproducibility and hinder the validation and generalization of AI-based interventions across diverse clinical contexts.

The Findable, Accessible, Interoperable, and Reusable (FAIR) guiding principles have emerged as a foundational framework for modern data management and open science practices [9]. First introduced in 2016 by Wilkinson et al. [9], these principles have been widely adopted across diverse scientific domains including agriculture, health, physics, computer science and computational biology as efficient data management framework [10–14]. In the context of medical science, the FAIR principles provide a robust framework for ensuring that research artifacts such as datasets and models are systematically organized, transparent, and readily available for validation, replication, and further exploration [15, 16]. Their applicability has been widely documented in the literature, extending to digital resources such as software [17], Machine Learning (ML) models [18, 19], and scientific workflows [20]. In the context of AI driven clinical trials, adherence to FAIR principles is particularly important. Similar to conventional medical interventions, AI interventions rely on high quality data, algorithmic transparency, and reproducibility across different settings.

Ensuring that AI models, training data (where relevant), and related metadata are FAIR can mitigate biases, enhance external validation and support long term sustainability of AI applications in healthcare [21].

In this study, we propose a novel and systematic approach to calibrate CONSORT-AI reporting guideline with FAIR principles. Rather than creating a separate framework, our goal is to provide methodological recommendations for integrating FAIR principles into CONSORT-AI, focusing on improving metadata reporting, accessibility of AI models and data, and refining reproducibility criteria. By incorporating these elements, both AI and guideline developers can play a pivotal role in strengthening the reporting standards for AI-driven clinical trials. While AI developers can enhance the FAIRness and transparency of their interventions regardless of current requirements, guideline developers are encouraged to reflect these practices in future updates. This shared responsibility ultimately improves the reproducibility and reliability of AI in clinical research.

## Methods

We used the calibration framework designed prior to this work (CALIFRAME) [22, 23] to calibrate CONSORT-AI with FAIR principles. CALIFRAME consists series of steps with predefined objectives and recommendations. The four main steps include i) Identification of reporting guideline and FAIR assessment tool, ii) Thematizing and mapping, iii) FAIR Calibration, and iv) Validation [22]. CALIFRAME is domain agnostic and can be applied to calibrate reporting guidelines with FAIR principles. We summarise below these four steps of the calibration exercise using the CALIFRAME framework as described in [22, 23].

### Stage 1: Identification of reporting guideline and FAIR assessment tool

The first stage of CALIFRAME involves identifying both an eligible reporting guideline (in the context of AI driven health care research) and a suitable FAIR assessment tool [22, 23]. We conducted a comprehensive and systematic literature review across several online databases, including PubMed, Web of Science, and specialized repositories such as the EQUATOR Network. Key criteria for selection included i) the guideline’s quality, ii) the rigor of its evidence-based recommendations, iii) its acceptance and use within the scientific community, and iv) its comprehensive coverage of essential reporting elements.

The first stage also involves identifying a suitable FAIR assessment tool to be used in the calibration process. The selection of this tool was based on its ability to provide a thorough and actionable evaluation of how well data and research outputs adhere to FAIR principles. The tool needed to be comprehensive, robust and user-friendly, with clear documentation and support for providing constructive feedback on data management practices.

### Stage 2: Thematizing and mapping

The second stage includes a systematic analysis of the selected reporting guideline to identify the core components and align them with the corresponding elements of the FAIR principles. We convened a series of virtual workshops that assembled an interdisciplinary group of experts, including researchers, specialists in FAIR principles, and developers of reporting guidelines. The workshop facilitated a thorough and multi-faceted examination of the identified reporting guideline and enabled a comprehensive evaluation of how well the guideline maps to the FAIR principles. The mapping process comprised a meticulous alignment of each theme, section, and subsection of the reporting guideline with the corresponding elements of the FAIR principles and FAIR indicators and metrics. It is important to note that this mapping task was not just a matching exercise; it included identifying gaps and areas where the guideline may not fully align with the FAIR principles.

### Stage 3: FAIR Calibration

In the third FAIR calibration stage, we conducted the second series of workshops to evaluate the alignment between the identified reporting guideline items and the FAIR principles.

Participants collaboratively examined areas of convergence and divergence between the two frameworks. This facilitated guided discussions aimed at exploring potential strategies for harmonization. The outcomes of these discussions were documented to inform the development of recommendations for enhancing FAIR-related components in reporting standards.

### Stage 4: Validation

During the validation stage, we verified that the recommendations and mappings made during the calibration process are not only theoretically sound but also practically applicable in enhancing the transparency, accessibility, and reusability of AI driven clinical trial data.

Thus, we assessed the effectiveness of the reporting guideline calibrated during the calibration workshops. We then reached out to the guideline developers for their feedback and critical review to further improve the calibrated reporting guideline items. We also shared the modified reporting guideline items with the RDA FAIR4ML-IG (FAIR for Machine Learning Interest Group) (https://www.rd-alliance.org/groups/fair-machine-learning-fair4ml-ig/activity/) and individually disseminated for experts including AI researchers and data stewards to gain a comprehensive feedback to further refine the reporting guideline.

## Results

The results from the calibration process are presented below with the details of milestones in each stage.

### Stage 1: Identification of reporting guideline and FAIR assessment tool

We conducted a systematic review to identify a reporting guideline to calibrate with FAIR principles, full detail can be found elsewhere [24]. We identified eleven (11) eligible reporting guidelines that were published before May 2024 and assessed the quality of each guidelines using the Appraisal of Guidelines, Research and Evaluation (AGREE-II) tool [25]. CONSORT-AI was selected for calibration due to its quality, as determined by our assessment, and its relatively wide endorsement within the research community [7]. It consists of 14 AI-specific items designed to complement the existing CONSORT 2010 core checklist, which together aim to improve the reporting of clinical trials involving AI interventions. Simultaneously, a detailed review of 14 FAIR assessment tools designed to assess FAIRness of all digital objects from the registry (https://fairassist.org/) were evaluated to select the most appropriate measure of FAIRness. FAIRMetrics [9] and RDA FAIR Maturity Model [26] were chosen for their systematic assessment of FAIRness with elaborated indicators and clear descriptions of how to do the assessment including the analysis. FAIRMetrics offers a set of measurable indicators for evaluating compliance with FAIR principles, while the RDA FAIR Maturity Model provides a community-driven, detailed methodology for conducting such assessments. Both tools include clear criteria and guidance on how to perform the evaluation and interpret the results.

### Stage 2: Thematizing and mapping the guideline

The mapping process commenced with the alignment of the CONSORT-AI items to each of the four principles of FAIR. This was subsequently refined by mapping to indicators in FAIR metrics and RDA FAIR maturity model.

All CONSORT-AI items mapped to one or more FAIR principles with most of the items being mapped to Findability and Reusability.

### FAIR mapping of the CONSORT-AI guideline

### Findability

All items in the ‘*Title and abstract’ and ‘Introduction’* sections of the CONSORT-AI are aligned with the Findability principle (Figure 1). The primary reason for mapping these items to Findability lies in their holistic descriptive nature, which serves as essential metadata for the study, AI intervention or dataset associated with the research. Additionally, items from the *‘Methods’ (3a,3b, CONSORT AI 4a (I, II)),* and *‘Results’ (14a,15,23)* sections were partially mapped to Findability principle. Although these items mapped to Findability principle, most of them mapped to specific metric FM_F2 (‘Data are described with rich metadata’) metrics of the FAIR Metrics [9] and RDA-F2-01M indicator (‘Rich metadata is provided to allow discovery’) of RDA FAIR indicators [27]. This means that other important aspects of Findability are overlooked. These include whether digital resources, such as datasets and AI models or source code have unique and persistent identifiers, and whether their metadata is indexed in searchable repositories in a way that enables discoverability.

**Figure 1:**
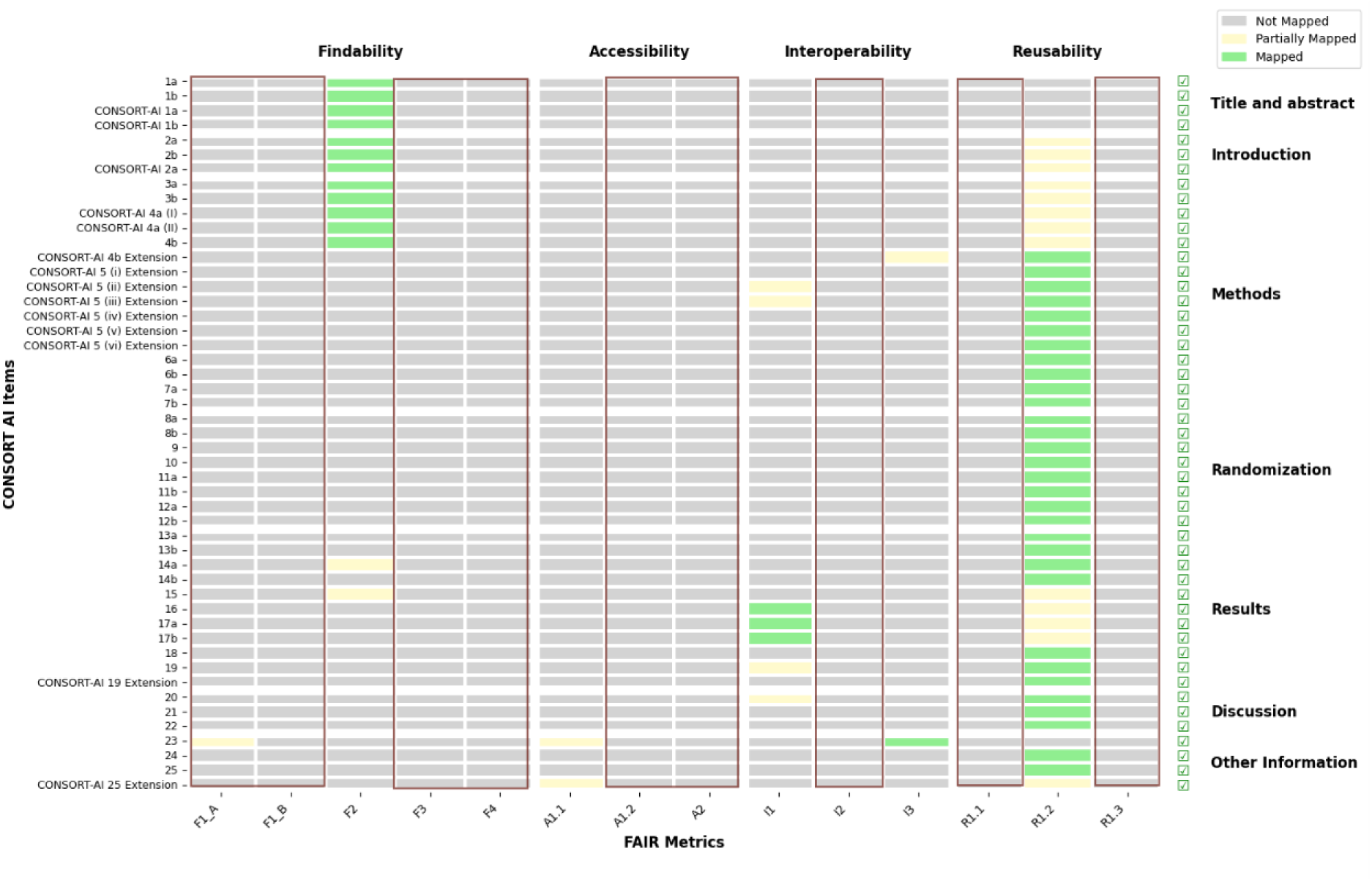
Mapping of CONSORT-AI reporting guideline to the FAIR metrics. The green checkbox on the right indicate that all CONSORT-AI items are mapped to one or more FAIR metrics and the light brown box indicate FAIR Metrics that are not addressed in CONSORT AI reporting guideline.

### Accessibility

With regard to Accessibility, only two items CONSORT-AI (Item 23 and ‘CONSORT-AI 25 Extension’) mapped to the Accessibility principle specifically FM-A1.1 (Access Protocol) and RDA-A1-01M (Metadata contains information to enable the user to get access to the data) from FAIR metrics and RDA FAIR indicators respectively. The explanation for item CONSORT-AI 25 Extension states that “*The trial report should make it clear whether and how the AI intervention and/or its code can be accessed or re-used. This should include details about the license and any restrictions to access*.”[6].

This item serves as an addendum to the CONSORT guideline specifically tailored to clinical trials with AI interventions, and presents a highly relevant reporting requirement for AI intervention. Moreover, it encompasses several aspects of FAIR principles condensed into a single item. For example, the item covers “*whether the code can be accessed or not”* (Accessibility), “*how the code/intervention can be accessed”* (Accessibility), “*whether the code/intervention is reusable”* (Reusability), how the *code/intervention is reused* (Reusability), “*licencing information”* (Reusability) and, “*access restriction”* (Accessibility). The condensation of multiple aspects of the FAIR principles into a single item, as seen in CONSORT-AI 25 Extension, poses several challenges for practical applicability. Firstly, the complexity of this item may lead to ambiguity in interpretation. Researchers, AI developers and practitioners may struggle to discern the specific requirements related to accessibility and reusability, which could result in inconsistent reporting practices. Secondly, the overlapping nature of the criteria may lead to incomplete compliance. For example, if a researcher focuses on one aspect such as providing licensing information, they may inadvertently neglect other critical components, such as detailing access restrictions or the specific methods for reusing the code. This could result in a situation where the AI intervention is technically accessible but lacks the necessary information for effective reuse. While we recognize the need to keep reporting items to a minimal in order to maintain applicability, this should not come at the expense of clarity and detailed reporting.

### Interoperability

Items in the ‘Intervention’ subsection (CONSORT-AI 4b Extension, CONSORT-AI 5 (II-III)), 16,17a,17b, 19 and item 23, were mapped to the Interoperability principle FM-I1 and FM-I3 from FAIR metrics, RDA-I1-01M and RDA-I3-01M from RDA FAIR indicators. The reason for mapping these items to Interoperability principle is that they focus on the detail descriptions (metadata) of the AI intervention used in the study, including version, and the details of pre-processing and other model related information.

Although there is an implicit expectation that researchers use standardized, machine-readable knowledge representations such as controlled vocabularies or persistent identifiers, it is not typical for researchers to adhere to these expectations unless explicitly requested to do so.

Researchers often operate under specific guidelines and frameworks that dictate what information is necessary for their reports and how it is reported. When elements such as controlled vocabularies or detailed metadata are not explicitly required by the reporting guidelines, researchers may not prioritize including them. This oversight can lead to gaps in the reproducibility of research.

### Reusability

Several items were mapped to Reusability principle specifically FM-R1.2 (detailed provenance) and RDA-R1-01M (Plurality of accurate and relevant attributes are provided to allow reuse). Provenance encompasses two primary type of information, as described by

Wilkinson et al. (2016): ‘who/what/when data/digital resource is produced’ and ‘why/how it was produced’ [9]. Additionally, CONSORT-AI 25 Extension mapped with Reusability principle due to its inclusion of the statement regarding licencing information (FM-R1.1).

The mapping process highlighted that all the additional items on CONSORT-AI guideline correspond to one or more of the FAIR principles, suggesting that these addenda are designed, either directly or indirectly to enhance transparency, reproducibility and FAIRness.

One of the challenges in mapping the CONSORT-AI reporting guideline with FAIR principles is ambiguity in terminology. For example: *CONSORT-AI 5(i) Extension* item (“State which version of the AI algorithm was used”) can be considered ambiguous. The term "algorithm" is often used to refer to the underlying mathematical model or method, while "version" could imply different iterations or updates of that algorithm. If the context does not clarify whether it refers to the specific implementation, the software version, or the overall AI system, it may lead to confusion. However, the explanation explicitly states that it refers to the version of the "AI system," not the "algorithm." It is important to distinguish between these terms, as they are often used interchangeably, leading to confusion.

### Stage 3: FAIR Calibration

From the mapping exercise, we identified elements of FAIR principles that are not represented in the CONSORT-AI reporting guideline, which if considered, could contribute to improving the transparency, reproducibility, and FAIRness of clinical trials with AI interventions. The calibration process attempts to fill this gap by proposing adjustments that align with FAIR principles, such as enhancing accessibility and promoting reusability of trial outcomes. These adjustments aim to bridge the gap between current reporting standards and the evolving needs of AI-driven clinical research, ultimately fostering greater trust and utility in AI interventions within the healthcare domain.

Table 1 presents the list of unmapped FAIR principle elements and the recommended calibration approach.

**Table 1:** Recommendations of FAIR calibration for CONSORT-AI reporting guideline.

| FAIR principles not addressed in CONSORT AI reporting guideline |  | Recommended calibration |
| --- | --- | --- |
| FAIR Metrics | RDA FAIR Indicator |  |
| F1A (Identifier Uniqueness) | RDA-F1-01M***, RDA-F1-01D***, RDA-F1-02M***, RDA-F1-02D*** | This principle requires digital object identifiers or other globally unique and persistent identifier for the AI intervention (source code) and the clinical trial dataset associated with the clinical trial.<br>**Recommendation:** Mandate the inclusion of these identifiers in the CONSORT-AI guideline. |
| F1B (Identifier persistence) |  |  |
| F3 (metadata clearly and explicitly include the identifier of the data it describes) | RDA-F3-01M*** | Make sure the identifier is explicitly included in the study and the trial report.<br>**Recommendation:** Introduce a requirement in CONSORT-AI to provide metadata that explicitly includes the identifiers linking to the underlying AI intervention and trial dataset and ensure the linkage is machine readable. |
| F4 (Indexed in a searchable resource) | RDA-F4-01M*** | Make sure that the AI intervention and the trial dataset (if applicable) is indexed in searchable resource.<br>**Recommendation:** Encourage registration of the AI intervention and the trial dataset (if applicable) metadata in searchable repositories or catalogues. Specify preferred repositories for the trial dataset and the AI intervention in CONSORT-AI. |
| A1.1 (Standardised Access Protocol) | RDA-A1-01M**, RDA-A1-02M***, RDA-A1-02D***, RDA-A1-03M***, RDA-A1-03D***, RDA-A1-04M***, RDA-A1-04D***, RDA-A1-05D**, RDA-A1.1-01M***, RDA-A1.1-01D** | Split CONSORT-AI 25 Extension, - State if the AI model/software/source-code is accessible. - If accessible, describe the access protocol (how to access). **Recommendation:** Require description of standardized protocols (APIs) for accessing trial data and AI models. Include guidelines for documenting these access methods. |
| A1.2 (Access authorization) | RDA-A1.2-01D* | **Recommendation:** split CONSORT-AI 25 Extension - Describe any restrictions with respect to the AI intervention/source-code access. |
| A2 (metadata are accessible, even when the data are no longer available) | RDA-A2-01M*** | Make sure that metadata is guaranteed to remain available even after the AI intervention or dataset is no longer available.<br>**Recommendation:** Introduce a requirement in CONSORT-AI to ensure metadata remains accessible even when full data or AI intervention cannot be shared. Provide guidelines for creating publicly accessible metadata for restricted datasets or AI models. |
| I2 (Use FAIR Vocabularies) | RDA-I2-01M**, RDA-I2-01D* | Annotate metadata with controlled vocabularies or ontologies.<br>**Recommendation:** Encourage the use of standardized vocabularies for describing trial data and AI model metadata. Recommend appropriate ontologies for AI specific concepts in CONSORT-AI. |
| R1.1 (Detailed Licence information) | R1.1-01M***, R1.1-02M**, R1.1-03M** | Make sure appropriate licence is provided associated with the trial dataset and AI intervention.<br>**Recommendation:** Encourage specifications of clear usage licences for all datasets and AI interventions. Provide guidance on selecting appropriate licences for different use cases in CONSORT-AI. |
| R1.3 (Meets Community Standards) | RDA-R1.3-01M***, RDA-R1.3-01D***, RDA-R1.3-02M***, RDA-R1.3-02D** | The metadata documentation should be structured in a community standard format.<br>**Recommendation:** Introduce a requirement to assess and report the compliance with community standard in CONSORT-AI. |

*RDA FAIR Indicators priority (\*\*\* Essential, \*\* Important, \*Useful)*

### F1A (Identifier Uniqueness) and F1B (Identifier persistence) Explanation

The F1A and F1B indicators emphasizes the necessity of assigning persistent identifiers (PIDs) to digital resources, such as AI models, software source code, and clinical trial datasets [9]. This principle is further elaborated by the RDA-F1-01M, RDA-F1-01D, RDA- F1-02M, and RDA-F1-02D indicators [27], which provide a structured framework for implementing identifier uniqueness. The recommended calibration for this principle mandates the inclusion of digital object identifiers (DOIs) or other globally unique and persistent identifiers for both the AI model/software/source code and the clinical trial dataset. This recommendation can be justified as follows:

*Traceability and Reproducibility:* Unique identifiers enable precise referencing of AI models and datasets, which is critical for reproducibility in scientific research [28]. Without such identifiers, it becomes challenging to trace the exact version of an AI model or dataset used in a clinical trial, leading to potential ambiguities and errors in replication studies.

*Credit and Attribution:* Unique identifiers ensure proper attribution to the creators of AI models and datasets, fostering a culture of recognition and accountability in the scientific community [29]. This is particularly important in the context of collaborative research and open science (OS) initiatives.

*Long-Term Accessibility:* Clinical trial data and AI models often have long term relevance, particularly in healthcare, where longitudinal studies and follow-up analyses are common. Persistent identifiers ensure that these resources remain accessible to future researchers.

*Avoidance of Link Rot:* In the absence of persistent identifiers, digital objects are susceptible to "link rot," where URLs or other locators become invalid over time [30]. This can render valuable research resources inaccessible, undermining the integrity of the scientific record. *Archival and Preservation:* Persistent identifiers are typically linked to reliable archival and preservation systems, which store digital objects securely and protect them from data loss or corruption [31]. This is particularly important for clinical trial datasets, which may be subject to regulatory requirements for data retention.

Mandating the use of globally unique and persistent identifiers for AI models and clinical trial datasets in the CONSORT-AI guideline enhances traceability, reproducibility, and long-term accessibility.

However, implementing this might be challenging. Linking to a code repository or dataset does not guarantee that the content will remain unchanged. Therefore, successful implementation of unique and persistent identifiers requires a robust system to generate, manage, and verify these identifiers, verifying their uniqueness, persistence, and provenance over time.

To address these challenges, the following steps are recommended:

1. *Adopt Established Identifier Systems:* Leverage existing systems like DataCite [32] or Crossref [33] for generating DOIs for datasets and AI models. These systems already provide globally unique and persistent identifiers and are widely used in the research community.
2. *Version Control for AI Models:* Require the use of version control systems (e.g., Git) for AI model source code (if applicable), with each version assigned a unique identifier. Platforms like GitHub, GitLab, or Zenodo can be used to archive and link specific versions of the code.
3. *Provenance Tracking:* Implement metadata standards (e.g., W3C PROV, PROV-O (Provenance ontology)) [34] to track the provenance of datasets and models, improving transparency and reproducibility.
4. *Incentivize Compliance:* Provide guidelines and tools to help researchers comply with identifier requirements. Funding agencies and journals could mandate compliance as a condition for publication or grant approval.

### Recommended changes in CONSORT-AI

Additional item: ***CONSORT-AI 5 (vii)****: Provide a unique and persistence identifier redirecting to the AI intervention used in the trial*.

Additional item: ***Item 13c:*** *Provide a unique and persistence identifier redirecting to the clinical trial dataset*.

### F3 (metadata clearly and explicitly include the identifier of the data it describes) Explanation

The FAIR principle F3 (Metadata clearly and explicitly include the identifier of the data it describes), supported by RDA-F3-01M, emphasizes embedding explicit identifiers in metadata to ensure traceability of AI interventions and clinical trial datasets. The recommended calibration mandates that metadata explicitly link to identifiers of the AI intervention and trial dataset. By requiring explicit, machine-readable metadata linking, this calibration strengthens transparency, and reproducibility of AI in clinical research. If the CONSORT-AI 5(vii) and 13c are properly incorporated in the reporting guideline, it will automatically resolve the machine readability and explicit inclusion of identifier.

### F4 (Indexed in a searchable resource) Explanation

The recommendation to address F4 (Indexed in a searchable resource) and RDA-F4-01M highlights the importance of making sure that AI models/interventions and trial datasets are indexed in searchable repositories [9]. The CONSORT AI reporting guideline does not explicitly require AI models or trial datasets to be indexed in searchable resources. This oversight limits the ability of researchers, clinicians, and policymakers to discover and reuse these resources, also hindering transparency and reproducibility. Without indexing, AI models and datasets remain isolated, decreasing their potential to contribute to broader scientific knowledge or to be validated in different contexts.

The recommendation proposes mandating the registration of AI interventions and trial datasets in searchable repositories. To standardize this process, specific repositories should be recommended. For example: For trial datasets: Repositories like ClinicalTrials.gov [35], Dryad [36], EASY [37], or Zenodo [38] can be used to register and share trial datasets if applicable. For AI models/Interventions: Platforms like ModelHub [39], HuggingFace [40], or ModelZoo [41] can serve as searchable catalogs for AI models.

### Recommended change in CONSORT-AI

The recommended changes in CONSORT-AI involve explicitly incorporating a requirement for the registration and indexing of AI models/interventions and trial datasets in searchable repositories.

Additional item: ***Item 23a***: *Provide referencing and indexing information of the AI intervention*.

Additional item: ***Item 13d***: *Provide referencing and indexing information of the trial dataset*.

### A1.1 (Access Protocol) and A1.2 (Access authorization) Explanation

The FAIR Metric A1.1 and A1.2 emphasize the importance of standardized access protocols and authorization mechanisms to facilitate transparent, efficient, and reproducible research [9]. They provide granular guidance on how metadata and data should be documented and accessed, so that both human and machine users can retrieve and utilize resources effectively [27].

The CONSORT-AI 25 Extension acknowledges the importance of accessibility by requiring authors to specify whether the AI intervention and/or its source code used in clinical trials are accessible. However, it lacks detailed guidance on how this accessibility should be documented and operationalized. To address this gap, we recommend to expand CONSORT-AI 25 to include explicit documentation of the access protocols and conditions.

### Recommended change in CONSORT-AI

The recommended changes in the CONSORT-AI include:

Additional item: ***CONSORT-AI 25 Extension (i):*** *Describe the standardized protocol used to access the AI intervention and/or source code and trial dataset (e.g., HTTP, FTP, API)*.

Additional item: ***CONSORT-AI 25 Extension (ii):*** *Describe whether the AI intervention and trial dataset can be accessed manually or automatically*.

Additional item*: **CONSORT-AI 25 Extension (iii):** Specify whether the access is free or restricted. If restricted, describe the access conditions (describe the access authorization process, including required credentials, user roles, and governance structures for approving access requests.)*.

### A2 (metadata are accessible, even when the data are no longer available) Explanation

Guaranteeing the long-term accessibility of metadata, even when the associated data or AI models are no longer available, is a core element of accessibility [42]. This is particularly important in clinical research, where AI models and trial datasets may be restricted due to ethical, legal, or proprietary concerns. Metadata plays a central role in documenting key attributes of models and trial datasets that makes them discoverable and interpretable even when direct access is not possible.

### Recommended Change in CONSORT-AI

To effectively implement this calibration, the CONSORT-AI guideline should explicitly integrate provisions for sustainable metadata preservation, ensuring relevant contextual information remains accessible even when datasets or models cannot be fully shared. This approach recognizes that metadata stewardship is typically managed through specialized registries and repositories, systems operating beyond individual researchers’ control.

*Additional Item*: ***CONSORT-AI 5(viii)****: Report whether the repository or platform hosting the AI intervention includes a policy that guarantee the long-term preservation and accessibility of associated metadata, even after the model itself becomes unavailable*.

Additional item: ***Item 13e:*** *Report whether the repository or platform responsible for the dataset provides a policy or mechanism to preserve and maintain access to metadata related to the dataset, even after the dataset itself becomes unavailable*.

### I2 (Use FAIR Vocabularies) Explanation

The FAIR principle I2 (Use FAIR Vocabularies) focus on the necessity of using standardized, shared, and machine-readable vocabularies to improve interoperability between datasets, metadata, and AI models [43]. The corresponding RDA Metric (RDA-I2-01M) underscores that controlled vocabularies, ontologies, and standardized terminologies must be used to describe AI interventions and trial datasets to ensure consistency and semantic clarity in clinical AI reporting [44]. In its current state, the CONSORT-AI guideline does not explicitly require the use of FAIR-aligned vocabularies for describing AI interventions, datasets, and study parameters [6]. The lack of standardized terminology in reporting AI-driven clinical trials creates inconsistencies that hinder data integration, reuse, and automation in systematic reviews and meta-analyses [45]. In general, the absence of shared vocabularies increases the risk of misinterpretation, reduces comparability across studies, and limits seamless data exchange.

### Recommended Change in CONSORT-AI

The recommended changes involve explicitly incorporating a requirement in CONSORT-AI to mandate the use of standardized vocabularies and ontologies for AI interventions, datasets, and study metadata.

Additional Item: ***CONSORT-AI 5(ix)****: Specify any standardized vocabulary, ontology, or terminology used to describe the AI system or its computational context. Where applicable, AI-relevant ontologies such as the Artificial Intelligence Ontology (AIO)* [46]*, Software Ontology (SWO)* [47]*, or EDAM* [48] *could be used. If the AI intervention relies on standardized clinical input data (for example, coded with SNOMED CT* [49]*, LOINC* [50]*, OMOP* [51]*, Mondo Disease Ontology* [52]*), also specify those vocabularies used for data representation as appropriate for the study*.

*Additional Item: **Item 13f**: Specify the controlled vocabulary or ontology used to annotate metadata for the trial dataset. Examples might include FAIRsharing* [53]*, OBO Foundry* [54]*,or BioPortal* [55]*tailored to the dataset’s domain)*.

*Additional Item: **Item 23b**: Indicate whether metadata and dataset annotations follow FAIR-compliant ontologies to facilitate semantic interoperability. This could involve adherence to* schema.org [56]*, Dublin Core* [57]*, or HL7 FHIR* [58] *standards where relevant)*.

### R1.1 (Detailed Licence information) Explanation

This principle underscores the necessity of explicitly specifying licensing terms for AI models, datasets, and metadata to clarify reuse conditions and address both legal and ethical considerations. The corresponding RDA Metric (RDA-R1.1-01M) requires that licensing information be machine-readable and sufficiently detailed to guide potential users regarding permitted and restricted uses [27]. Clear licensing is essential not only to facilitate legal compliance but also to support collaborative research and data sharing, which are integral to enhancing the reproducibility and scientific impact of clinical trials.

In its current version, the CONSORT-AI guideline item ‘CONSORT AI 25 Extension’ requires a statement regarding access and restriction of AI intervention [6] but does not mandate separate, explicit licensing information for AI models, datasets, or metadata which are distinct resources. This omission can lead to uncertainty regarding the legal and ethical reuse of these resources, potentially limiting their modification, redistribution, and overall utility. To address this limitation, we recommend that CONSORT-AI explicitly incorporate licensing requirements.

### Recommended Change in CONSORT-AI

Additional Item: ***CONSORT-AI 25(i):*** *specify the license under which the AI intervention and its associated metadata are made available (e.g., CC-BY* [59]*, MIT* [60] *,GPL* [61] *proprietary or an alternative licence appropriate to the context)*.

Additional Item: ***CONSORT-AI 25(ii)***: *specify the license governing access and reuse of the trial dataset, including any restrictions or conditions (e.g., CC0* [59]*, Open Data Commons* [62]*, Data Use Agreements* [63]).

Additional Item: ***CONSORT-AI 25(iii):*** *state that licensing information is provided in a machine-readable format, allowing automated processing and indexing*.

### R1.3 (Meets Community Standards) Explanation

The R1.3 (Meets Community Standards) principle emphasizes the need for datasets, metadata, and AI models to adhere to recognized community standards to ensure widespread usability, integration, and trustworthiness [64]. The corresponding RDA Metric (RDA-R1.3- 01M) specifies that data and metadata should conform to domain-specific best practices, promoting consistency and quality in scientific reporting. With this regard, CONSORT-AI guideline does not explicitly mandate adherence to established community standards for AI model and dataset documentation. This may lead to inconsistencies in reporting, reduced interoperability, and challenges in validating AI-driven clinical research across different studies and institutions.

To address this, we recommend incorporating a requirement for adherence to recognized community standards in AI-driven clinical trials.

### Recommended Change in CONSORT-AI

Additional Item***: CONSORT-AI 5(x):*** *Indicate whether the AI intervention follows community-accepted standards for model development, validation, and documentation (e.g., ISO/IEC 23053* [65]*, ISO/IEC 2382-37* [65]*, TRACE* [66], *MLOps* [67] *or other appropriate community standard)*.

Additional Item: ***Item 13g:*** specify the community standards followed for structuring and annotating trial datasets (e.g., HL7 FHIR [58], OMOP Common Data Model [68] or other).

Additional Item: ***CONSORT-AI 23c***: *State weather metadata and dataset annotations comply with domain-specific community standards to enhance interoperability and reliability*.

### Stage 4: Validation

TBA based on the next iteration of feedback from CONSORT-AI developers + FAIR4ML IG

## Discussion

Our FAIR-driven calibration of CONSORT-AI identified opportunities to enhance reproducibility in AI trial reporting. Specifically, we observed that certain elements essential for comprehensive FAIR compliance are not yet explicitly addressed. For instance, CONSORT-AI does not currently include items requiring persistent global identifiers for datasets or models, rich metadata connecting data and code to the trial, or machine-readable licensing and repository indexing. These findings align with earlier reviews that noted limited reporting of algorithmic details and code availability in AI trial publications [69, 70].

In our work, we show how FAIR principles directly target these known challenges. Embedding FAIR criteria into CONSORT-AI contributes to more concrete guidance on data/code sharing and provenance. As Haibe-Kains et al. argue, “computational reproducibility is indispensable for high-quality AI applications”, and without code the only option is to “replicate methods from textual description,” which is often infeasible [71].

Our analysis generated nine targeted recommendations aimed at enhancing the reproducibility and FAIR compliance of AI trial reporting. While the full set of proposals is detailed in the results, several cross-cutting priorities emerge. These include the implementation of persistent identifiers for digital resources, the provision of structured and sustainable metadata, and the indexing of data and code in accessible public repositories.

Additional recommendations emphasize the use of machine-readable licenses, alignment with domain-specific standards, and adoption of controlled vocabularies. Collectively, these measures are intended to support long-term accessibility, interoperability, and reuse of digital artifacts associated with AI trials.

By operationalizing FAIR at the guideline level, we turn abstract openness goals into actionable checklist items. As our prior framework summary notes, integrating FAIR with reporting standards “bridges the gap between FAIR metrics and current reporting,” [72]. In this way, our calibration builds on and extends existing efforts by systematically translating established open-science imperatives into improved trial reporting.

## Limitations

This qualitative calibration has limitations. First, our mapping from FAIR principles to CONSORT-AI items was based on expert workshops and thematic analysis, which inevitably involves interpretation. Different stakeholders (e.g. clinicians, data scientists, ethicists) might map certain FAIR elements differently or prioritize others. We mitigated this with multidisciplinary feedback, but subjective biases may remain. Second, we focused solely on CONSORT-AI. Although CONSORT-AI is a prominent guideline, it represents only one part of the AI-trial ecosystem. Other documents (e.g. SPIRIT-AI for protocols, TRIPOD-AI for predictive models) may present unique challenges or opportunities not captured here. Third, our work to date has not included empirical testing, the proposed checklist items and workflows must be validated. We have solicited input from the CONSORT-AI group and the RDA FAIR4ML community, but formal consensus and adoption will depend on future guideline updates and institutional timelines.

## Practical implications

The calibrated CONSORT-AI has practical implications across the research ecosystem.

*Guideline developers*: Our work offers a clear methodology to update existing or new guidelines with data-stewardship principles. Guideline developers can incorporate the recommended items in the checklist or separate elaboration document as appropriate (e.g. in a future CONSORT-AI revision) to advance transparency without reinventing whole frameworks. Finding the right balance between comprehensiveness and usability will require careful iterative refinement.

*Researchers and trialists*: The FAIR-enhanced checklist provides concrete expectations for reporting AI trials. By planning data management and code sharing early, investigators can ensure compliance and reduce ambiguity (for example, by pre-registering persistent identifiers or metadata standards).

*Journals and funders*: Current endorsement of CONSORT-AI is low and even fewer require FAIR data practices. By demanding adherence to calibrated guidelines, journals can improve review quality and regulatory compliance.

## Conclusion

This work introduces a FAIR calibration of the CONSORT-AI reporting guideline, formally linking rigorous trial reporting with modern data stewardship practices. By encouraging authors to make trial data and AI models Findable, Accessible, Interoperable, and Reusable, the calibrated guideline can improve the reproducibility, transparency, and long-term value of clinical AI research. In effect, it elevates the role of data and code sharing to the same level as other trial design elements. This approach has the potential to strengthen the evidentiary basis for AI interventions. Looking ahead, stakeholder workshops and pilot testing will be important to refine the calibrated guideline.

## Data Availability

All data produced in the present work are contained in the manuscript

